# Multimodal and Quantitative Analysis of the Epileptogenic Zone in the Pre-Surgical Evaluation of Drug-Resistant Focal Epilepsy

**DOI:** 10.1101/2024.07.11.24310242

**Authors:** Hamid Karimi-Rouzbahani, Simon Vogrin, Miao Cao, Chris Plummer, Aileen McGonigal

**Author notes:** Joint first author (Hamid Karimi-Rouzbahani, Simon Vogrin). Joint last author (Chris Plummer, Aileen McGonigal). Correspondence to: Hamid Karimi-Rouzbahani.

## Abstract

Surgical resection for epilepsy often fails due to incomplete Epileptogenic Zone (EZ) localization from standard electroencephalography (EEG), stereo-EEG (SEEG), and Magnetic Resonance Imaging (MRI). Subjective interpretation based on interictal, or ictal recordings limits conventional EZ localization. This study employs multimodal analysis using high-density-EEG (HDEEG), Magnetoencephalography (MEG), functional-MRI (fMRI), and SEEG to overcome these limitations in a patient with drug-resistant MRI-negative focal epilepsy. A teenage boy with drug-resistant epilepsy underwent evaluation. HDEEG, MEG, fMRI, and SEEG were used, with a novel HDEEG-cap facilitating simultaneous EEG-MEG and EEG-fMRI recordings. Electrical and magnetic source imaging were performed, and fMRI data were analysed for homogenous regions. SEEG analysis involved spike detection, spike timing analysis, ictal fast activity quantification, and Granger-based connectivity analysis. Non-invasive sessions revealed consistent interictal source imaging results identifying the EZ in the right anterior cingulate cortex. EEG-fMRI highlighted broader activation in the right cingulate cortex. SEEG analysis localized spikes and fast activity in the right anterior and posterior cingulate gyri. Multi-modal analysis suggested the EZ in the right frontal lobe, primarily involving the anterior and mid-cingulate cortices. Multi-modal non-invasive analyses can optimise SEEG implantation and surgical decision-making. Invasive analyses corroborated non-invasive findings, emphasising the importance of individual-case quantitative analysis across modalities in complex epilepsy cases.

## Introduction

There are over 50 million people with epilepsy worldwide [14]. Anti-seizure medications (ASMs) cannot adequately control the disorder in at least 30% of cases [34]. If the epilepsy is considered focal (seizures arising from part of one hemisphere) [15], those with drug-resistant focal epilepsy may undergo presurgical evaluation to detect those areas that are critical for the generation of seizure activity. These areas have been variably termed the epileptogenic zone (EZ) or the epileptogenic network. The goal of surgery is to laser-ablate, resect, or disconnect the EZ by sacrificing the least amount of brain tissue to render the patient seizure free while avoiding the creation of a functional deficit.

The inherent complexity of the epileptogenic zone (EZ) and our limited ability to reliably and accurately localise it contributes to the failure of the surgical approach to achieve long-term seizure freedom for these patients [40]. Putting aside any conceptual considerations of how best to define the EZ [39,46], this problem is under-pinned by two key factors - the inability of any single imaging modality to routinely define the EZ and the subjectivity that is routinely brought to the analysis and interpretation of the recorded data. For example, standard magnetic resonance imaging (MRI), scalp electroencephalography (EEG) and stereo-EEG (SEEG) provide incomplete, albeit somewhat complementary, views of the brain. Specifically, functional MRI (fMRI) provides images of the brain at high spatial resolution (mm-scale), while lacking the necessary temporal resolution to delineate the rapidly changing brain activities typical of epileptiform discharges. MRI methods generally record resting state (interictal) activity between seizures (spikes and sharps). Standard low-density scalp EEG, on the other hand, provides sufficient temporal resolution (ms-scale) and allows study of both seizure (ictal) and interictal periods. However, its lower spatial resolution (cm-scale), which is affected by the complex and often imperfectly modelled electromagnetic properties of the tissues between the brain and the recording electrodes (primarily the skull) limits precise localisation of signal sources. The use of more recording electrodes with High density EEG (HDEEG) (at least 64 electrodes that includes a minimum three-pair inferior temporal electrode chain) combined with realistic MRI-individualised head models for electrical source imaging (ESI) partly alleviates this limitation (to at best 4-6cm^2^). While less available, magnetoencephalography (MEG) for magnetic source imaging (MSI), is less influenced by tissue interfaces and has a better spatial resolution (to at best 1-3cm^2^). However, like MRI-based techniques, the requirement for patients to remain still for both HDEEG and MEG usually limits their data capture to interictal events. In patients in whom invasive recording is deemed necessary for presurgical evaluation, SEEG can potentially overcome these issues with its high temporal and spatial resolution of anatomical correlates of interictal and ictal activity in the context of seizure clinical characteristics (semiology) captured during video scalp EEG monitoring. The additional advantage of SEEG over scalp EEG is that SEEG has minimal artefact from muscle contraction and movement during the ictal recording that can significantly distort the neural signals recorded during seizures. The downside of SEEG is its under-sampling of the brain - the number and positioning of implanted electrodes is limited by safety and technical measures such that only 5-10% of the brain is routinely sampled. The implication is that the SEEG trajectories are determined by EZ hypotheses formulated from the clinical teams’ interpretation of all the available non-invasively acquired data and the clinical presentation. The more experienced the clinical team and the higher the quality of the non-invasive datasets, then the higher the quality of the hypothesis for EZ localisation and the higher the likelihood that the true EZ will actually be sampled during the SEEG monitoring period. Therefore, these distinct recording modalities, in the context of expert interpretation, provide complementary views of the brain. As each of the above modalities (scalp EEG, MEG, SEEG, MRI and fMRI) provides specific benefits over one another, and based on the marked heterogeneity of epilepsy across individuals making the localisation difficult, one solution might be to utilise complementary strengths of multi-modal brain recording for clinical localisation of the EZ. This work evaluates the additional value of combining multiple brain recording data including high-density EEG (HDEEG), magnetoencephalography (MEG), fMRI and SEEG for the localisation of the EZ.

Another critical issue in EZ localisation is that conventional localisation of the EZ (and the potentially wider epileptogenic network) is generally reliant on subjective and qualitative interpretation of the available data. For example, structural MRI images are visually inspected for the detection of potential lesions and dysplasia. Hours of EEG and SEEG data are inspected to detect patient-specific epileptiform patterns of activity including established patterns such as low-voltage fast activity (LVFA) [36] in the ictal period or high-frequency oscillations (HFOs) and spike activity [9,50] during interictal periods. The level of expertise of the clinicians in their interpretation of data and the associated inter-rater variability can significantly impact the working hypothesis for the likely EZ localisation. Moreover, the complexity and the patient-specific nature of epileptiform brain activity can create challenges for the clinician to discriminate epileptiform activity from healthy brain activity [16]. To help overcome these issues, semi-automated and automated quantitative methods for the localisation of EZ has been developed for each modality separately.

We previously described approaches to the localisation of the EZ using HDEEG and MEG [4,5,42,51]. Specifically, we found that the earliest localisable interictal and ictal activity is a more accurate marker of the origin and extent of the EZ rather than later phase activity at either the mid-upswing or peak phases of these discharges. While the latter phases have been traditionally ear-marked as the most reliable time-point for source imaging solutions on account of the higher SNR characteristics, we found that this activity is commonly contaminated by the effects of cortico-cortical propagation for both interictal and ictal discharges. We also found that the combination of HDEEG and MEG carries complementary non-redundant source imaging information for non-invasive pre-surgical mapping of the likely EZ. A particular advantage of HDEEG based ESI is that its reproducibility can be assessed for solution stability between HDEEG-MEG and HDEEG-fMRI recordings across sequential sessions. That is, not only can the HDEEG inform the timing of the fMRI BOLD response, but it can also serve to define a second ESI result that can be cross-checked with that obtained from the HDEEG during the earlier HDEEG-MEG acquisition.

Despite the reduced temporal resolution of fMRI against EEG, MEG and SEEG, its high spatial resolution offers unique insights into secondary haemodynamic features correlated with neuronal activation related to interictal and ictal discharges. Approaches typically employed for fMRI include a priori model-driven analyses (such as general linear modelling, GLM, using canonical haemodynamic response functions), data-driven analyses (via Independent Component Analysis, ICA) and functional connectivity exploration (via Regional Homogeneity, ReHo metrics). The GLM approach is dependent upon simultaneously acquired EEG (EEG-informed fMRI), while ICA and ReHo approaches do not rely on co-acquired EEG data.

In SEEG, quantitative EZ localisation methods have been growing in the past two decades. These methods quantify patterns of epileptogenic activity either in the interictal period (e.g., spikes, sharps, HFOs, or complex signals, [32]) or ictal period (e.g., slow shifts and LVFA, [36]). Recent studies have localised the EZ by tracking epileptogenic patterns of activity from one area to another using connectivity techniques such as latency of spike activity [11,47] in the interictal period or connectivity metrics in the ictal period [18,19,38]. These connectivity approaches build upon previous studies that looked at individual brain areas to localise the EZ and are supported by growing evidence suggesting that focal epilepsy is a network disorder rather than a localised brain disorder [33,48,52]. We recently showed the additional information that connectivity measures provide to the elucidation of the EZ [32].

In light of the modality-specific limitations of EZ localisation and motivated by recent shifts towards quantitative analyses of epilepsy, this study uses a combination of HDEEG, MEG, fMRI and SEEG based on a sequence of novel analyses. We take advantage of each modality’s particular strength and show concordant multi-modal evidence for the location of the EZ in a patient with severe drug-resistant MRI-negative focal epilepsy, which in this complex case helped to refine SEEG implantation strategy and achieve satisfactory EZ localisation and ultimately treatment.

## Methods

### Case

A teenage boy with onset of epilepsy in childhood underwent presurgical evaluation for MRI-negative drug-resistant seizures. Despite two previous right prefrontal resections in another expert epilepsy centre in childhood (with evidence of focal cortical dysplasia type 2A on one resection), he continued to suffer daily (often 5-10 per day) hyperkinetic seizures, sometimes preceded by a sense of the heart racing or a sensation described by the patient as “a feeling in my heart and in my head”, and sometimes a feeling of being frightened. Objective seizure semiology was complex, characterised by partially retained awareness, vocalisation (where the patient could swear or shout “help”), pallor and abrupt, early onset of a hyperkinetic phase with rapid side-to-side pelvic and truncal movements while lying on the bed. Retained motor function of right arm was noted in some seizures (would grip the bedrail) and in about half of seizures he would turn to the prone position (via either the right or the left).

There was no clear-cut tonic posturing, though he would sometimes have the left arm in a flexed position. There was not a clearly emotional facial expression, though the content of the vocalisation could suggest an emotional experience. There was no incontinence and he never had secondary generalisation. Seizures were brief (<1 minute), appeared stereotyped in their motor sequence and had rapid recovery to baseline (no post-ictal motor or language deficit). In recorded seizures the patient was noted to be pale and sweaty during the hyperkinetic and post-ictal phase. He could feel nauseated and sometimes vomit at seizure offset. Interictal neurological exam was normal; neuropsychiatry and neuropsychologic testing showed low-average intelligence, reduced executive function and attentional deficits.

In terms of localising value of semiologic data, the presence of autonomic features (heart racing, pallor, sweating, nausea) suggested likely involvement of the central autonomic network (amygdala, anterior insula, anterior cingulate). The pattern of hyperkinetic motor behaviour suggested prefrontal and/or insular involvement at onset, with in particular turning to the prone position being suggestive of anterior or mid-cingulate involvement [41]. The very stereotyped pattern of seizures indicated a likely unifocal EZ, which was thought most likely to be within right frontal lobe, although independent or even primary EZ extra-frontal involvement of connected limbic structures (amygdala, insula) could not be excluded from the standard pre-surgical work-up. Indeed, interictal surface EEG during video-EEG monitoring showed sharp theta over right temporal and anterior suprasylvian regions. No discernible ictal EEG change was seen on video-EEG (22 seizures recorded). The SPECT was felt to be practically difficult (because of marked early hyperkinetic movements) but also not considered likely to bring clinically useful spatial resolution with regards to timing of injection since propagation can be expected to be extremely fast in such cases. It also seemed unlikely that SPECT would change the SEEG implantation strategy. We thus proceeded to additional investigations to aid in hypotheses for SEEG planning for this patient’s focal drug-resistant epilepsy. The patient has since undergone surgery (follow-up 6 months), with histopathology showing FCD 2A and marked improvement in seizure frequency and severity (Engel Class II).

Ethics approvals for this study were obtained from Institutional Review Board of Mater Misericordiae Ltd. Human Research Ethics Committee and Institutional Review Board of the Swinburne University of Technology. The patient consented to the multimodal investigation of his epilepsy at Mater Hospital and the Swinburne University of Technology. He also consented to the use of his data for research purposes and for publication.

### HDEEG and MEG

The patient underwent a series of non-invasive multimodal investigations across four stages (HDEEG-only, HDEEG-MEG, HDEEG-fMRI, structural MRI-only) sequentially performed during a single recording session at Swinburne University of Technology, (Melbourne, Australia). A 134 channel-HDEEG acquisition system (SynampRT™ EEG amplifier and Curry 9™Acquisition software, Compumedics Neuroscan, Melbourne) was employed at each stage. To control for the comparability of identified epileptiform brain activity across a similar baseline brain state for each recording stage, the patient and their treating team consented to one of two typical clinical provocations to enhance the yield of interictal discharges: minor sleep deprivation (2 hours less sleep), withholding of morning antiseizure medications until after testing completed (while under the constant clinical supervision of one of the authors (CP)). The combined duration of the sessions was limited to less than six hours. A novel 134 electrode HDEEG cap (10-10 system with 12 electrode inferior temporal coverage) constructed from non-magnetising hardware components was integrated for use with the MicroMaglink™ passive filters (Compumedics Neuroscan, Melbourne). The set-up minimised external RF noise contamination of the MEG and MRI scanner environments, permitting sequential recordings of (i) HDEEG-only in an electrically shielded room, (ii) HDEEG-MEG in a magnetically shielded room simultaneous with the 306 sensor MEG scanner (Triux, Elekta Oy), and (iii) HDEEG-fMRI session inside the MRI scanner (3T Prisma FIT, Siemens, Erlangen) without the need to reapply the HDEEG cap across these different recording environments prior to considerations for SEEG investigation. The reproducibility of ESI (EEG source imaging) was, therefore, based on HDEEG alone, HDEEG during MEG, and HDEEG during fMRI. ESI and MSI (MEG source imaging) results were also compared.

### HDEEG

HDEEG data were acquired at a sampling rate of 5 kHz (passband DC – 2kHz) across all stages (HDEEG-only, HDEEG-MEG, HDEEG-fMRI). HDEEG electrode positions, and PAN (pre-auricular, nasion) co-ordinates, along with samples of the cap surface, were digitised optical sensor tracker system (NDI Polaris Vicra®) in common space for MRI co-registration. Impedances were kept below 10 kOhms.

### MEG

The MEG data (102 magnetometers, 204 planar gradiometer) were sampled at 5000 Hz with anti-aliasing filter set at 1650 Hz. MEG head coils, HDEEG electrode positions, and PAN co-ordinates were re-digitized with an independent electromagnetic tracker (Polhemus Fastrak®). Bad channels for all MEG data were checked prior to applying temporal extension to signal source separation (tSSS) using Maxfilter® v2.2.15 (Elekta Oy) for interference suppression (correlation limit 0.98 and sliding window of 10 s). Independent 1-s interval clock triggers were captured on each system and used to synchronize HDEEG and MEG offline. The synchronisation was verified by comparing ECG channel signal phase from each independent modality. All HDEEG and MEG signals were reviewed and analysed using Curry 9 software. Applying a bandpass Hann-shaped FFT filter from 1 to 100 Hz (slope of 2 Hz, 50% transmission at 1 Hz; slope of −20 Hz, 50% transmission at 100 Hz), IEDs were manually identified by an experienced neurologist (C.P.) on HDEEG and MEG independent files using Curry 9™ (Compumedics Neuroscan, Hamburg). Independent-modality source localization (ESI, MSI) was performed. ESI and MSI were performed using (sLORETA) standardised low-resolution electromagnetic tomographic analysis (inverse model) and individualised MRI informed (BEM) boundary-element method (forward model) on averaged epileptiform discharges at different latency points (take-off, early-phase or earliest stable solution to explain 80% measured field, mid-phase or mid-upswing, late-phase or surface negative peak).

### HDEEG-fMRI

Simultaneous HDEEG-fMRI was acquired using a Multi-Echo Multi-Band (MEMB) sequence in a 3T Siemens PrismaFIT MRI scanner with a 32-channel head coil, with SynampRT amplifier synchronised to the 10MHz MRI clock signal. First, a high-resolution 3D T1-weighted magnetisation-prepared rapid acquisition with gradient echo (MPRAGE) anatomical image was acquired with TR/TE = 1900/2.26 ms, FOV = 24 cm, matrix size = 256 × 256 × 176, slice thickness = 1.0 mm, voxel size = 1.0 × 1.0 × 1.0 mm, and flip angle (FA) = 9°. The MEMB scan had the following parameters: TR/TE = 1392/10.6, 24.37, 38.14, 51.91, 65.68 ms (five echoes), FOV = 21.6 cm, matrix size = 86 × 86 with slice thickness = 2.5 mm (2.5 × 2.5 × 2.5 mm voxel size), 15 slices with multiband factor = 4 utilized in-plane acceleration (R) = 2 (total slices 60), FA = 90°, and partial Fourier factor = 0.75. Resting state HDEEG-fMRI scanning lasted for about 30 mins (1810 secs), or 1300 dynamic scan timepoints (6500 EPI volumes in total). For this acquisition, the patient was instructed to close their eyes, but remain relaxed and awake, to refrain from any motion aside from steady breathing throughout, and not think about anything in particular. Upon completion of functional scanning, the patient exited from the scanner and photographs verifying HDEEG cap head-alignment were captured immediately upon cap removal. An additional structural MRI-only stage, including a repeat of the earlier MPRAGE scan, was performed explicitly without HDEEG cap.

MEMB pre-processing and denoising: Three (24.37, 38.14 & 51.91 ms) of the five echoes datasets were independently processed for exploring BOLD dependence and robustness of potential EZ components, with each dataset independently registered to the initial cap MPRAGE image with optimization using boundary-based registration [20]. Respective echo datasets were independently denoised using a spatial ICA-based strategy for Automatic Removal of Motion Artefacts (ICA-AROMA) with automatic dimensionality estimation and non-aggressive denoising [13,43]. ICA-AROMA is an unsupervised data-driven technique based on the FMRIB Software Library (FSL) tool MELODIC [1] that removes motion-related (bad) components from the data, exploiting a small, robust set of features for classifiers without training. The denoised fMRI data from each echo were also separately analysed to assess features from both EEG-naive and EEG-informed BOLD (blood oxygen level dependent) effects. Candidate EZ regions from remaining components of a subsequent spatial-ICA of the denoised data were extracted, manually classified applying criteria adopted from a working paradigm [2]. Independently, the denoised but unsmoothed data was 0.01-0.1Hz band-passed and transformed to extract static Regional Homogeneity (ReHo) measures using FATCAT (Functional and Tractographic Connectivity Analysis [49]) software library available under AFNI (Analysis of Functional NeuroImages) software suite [7]. Dynamic ReHo time-series were constructed from sequential 20sec sliding windows, prior to decomposing the ICA components for estimating local connectivity of candidate EZ sources in MRI space.

Prior to clinical review of IEDs by author (C.P), MRI-gradient and pulse artefact-correction of HDEEG data were applied consecutively using an average artefact subtraction (AAS) approach with Curry 9™. A subsequent bandpass Hann-shaped FFT filter from 1 to 40 Hz (slope of 2 Hz, 50% transmission at 1 Hz; slope of −8 Hz, 50% transmission at 40 Hz) was applied to address cryogenic pump noise artefacts above this range, when marking event timings of candidate IED peaks, to generate a set of event regressors for GLM, using FEAT (FMRI Expert Analysis Tool, FMRIB Software Library [53]). These timings were also applied to compare the models against time-courses of EEG-naïve BOLD ICA.

### SEEG

#### Data acquisition

The target locations for SEEG electrodes were determined based on seizure semiology, the previous resections’ pathology data suggesting focal cortical dysplasia in the right frontal lobe, video-EEG data, ESI, MSI, and fMRI results. Accordingly, the working hypothesis was right frontal lobe epilepsy, with insular and temporal lobe sources as alternative EZ candidates, or as part of a more widespread EZ. Eleven electrodes were implanted on the right and three electrodes on the left hemisphere to rule out EZ being on the left. See the electrode labels and the area that each contact records from based on Hammers 95 brain atlas’s segmentation [23] in Supplementary Table 1 and the locations of the electrodes overlaid on the patient’s pre-implantation MRI in Figure 1.

**Figure 1.**
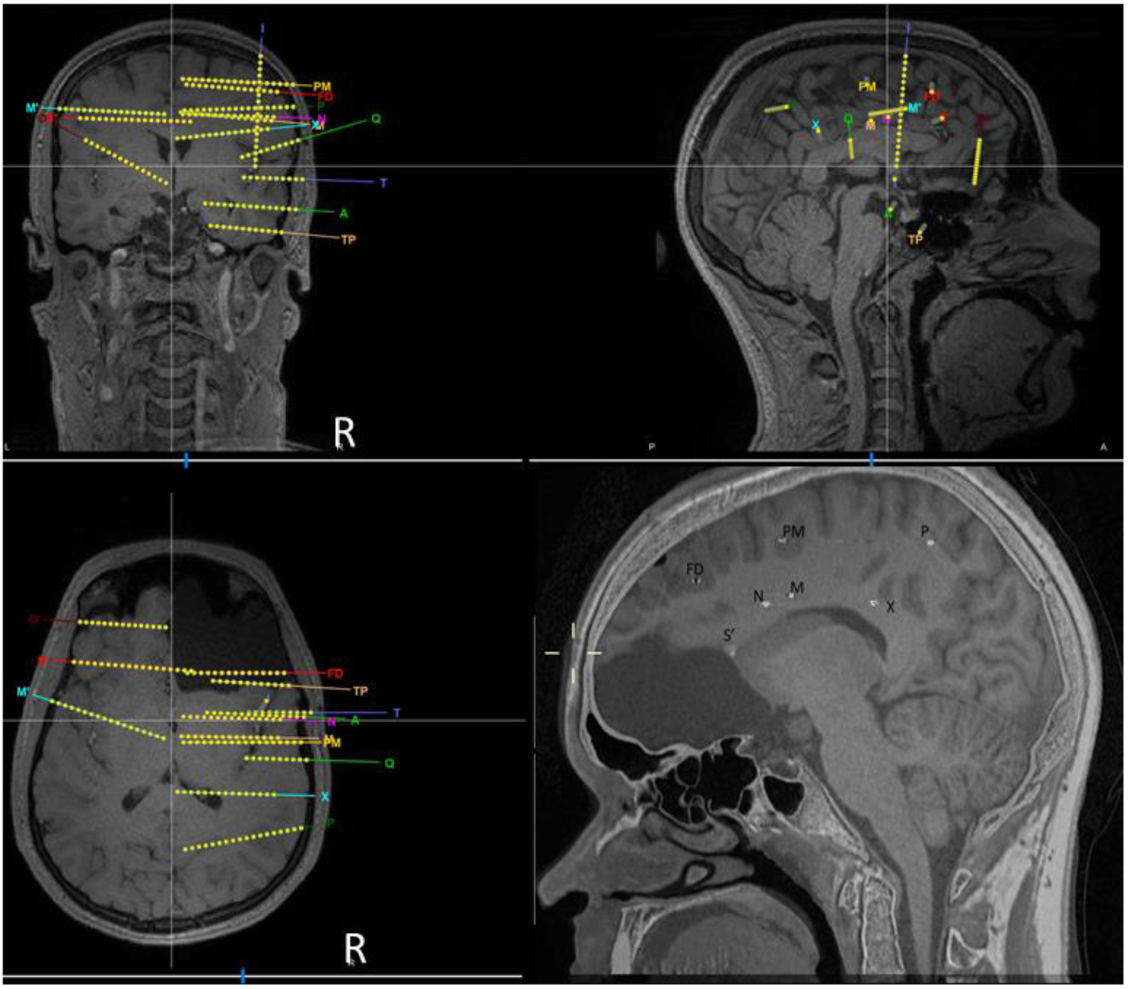
Reconstructed electrode and contact locations overlaid on the patient’s preimplantation MRI (top and left bottom panels). Right bottom panel shows the fused pre-implantation MRI and post-implantation CT to indicate the location of tip of the electrodes inserted through the right hemisphere and the S’ electrode inserted through the left hemisphere.

The patient underwent SEEG recording over 5 days, with partial ASM reduction on day 1 and 2, and reinstatement of usual doses on day 3. A total of 9 habitual seizures were recorded during the 5-day SEEG monitoring, all similar electro-clinically. These were spread throughout the recording period, with 1-2 spontaneous seizures occurring each day of recording from day 1-day 5.

The SEEG data was collected using a Nihon-Kohden EEG system, which recorded the signals at a sampling frequency of 1000 Hz in the frequency range above 0.08 Hz. Bad channels were excluded from analyses based on visual inspection. No other pre-processing analyses were performed on the data. The interictal data were epoched to include four 30-minute recordings, two on the 1^st^ day (>24 hours post anaesthesia) and two on the 4^th^ day of implantation in wake and sleep stages, respectively, at least 2 hours from any seizure. The reason for using 30 minutes of recording was a matter of practicality to ensure that the epoch was long enough to provide enough interictal spiking for analysis but not too long so that the patient’s mental and physical status would drastically change within the epoch. We chose to analyse both wake and sleep interictal states because of the known effect of sleep on interictal spike rate and electrical field [17]. We chose days 1 and 4 to evaluate both early and later stages of monitoring in the analysis. The ictal data was epoched from *30 to +30 seconds relative to the onset of each of the 9 recorded seizures determined visually by the initiation of electrographic seizure activity.

#### Interictal analyses

We detected spikes across all channels using an automatic spike detector in wake and sleep stages to localise the irritative zone. The automated spike detection algorithm detected HFOs (80-250 Hz) that co-occurred with interictal epileptiform spikes [6], which has been shown to be a more robust indicator of epileptogenic tissue compared to the detection of spikes alone [44]. The algorithm was run on the four 30-min time windows of the interictal recording – two on day 1 (wake and sleep) and two on day 4 (wake and sleep). The algorithm needed the user’s confirmation on every individual detected spike. To avoid subjective inputs, we accepted all the interictal spikes detected by the automatic detector upon making sure that they looked genuine and free of artefact.

Our second interictal analysis identified potential regions of spike initiation before propagation based on interictal spike co-occurrence and temporal progression across the SEEG contacts [24]. Specifically, we first selected four contacts in which we observed the maximum number of spikes and then checked the time delay between their co-occurring spikes (i.e., those which occurred within 1 s time delay from each other across contact pairs), to track potential spike propagation across the brain. The time of spike onsets were used in analyses. This method aligns with recent studies which used spike timing to localise the EZ [11,47].

#### Ictal analyses

We first generated a cortical “epileptogenicity map” of the potential seizure onset zone which quantifies the level of fast activity during seizures across areas sampled with electrode contacts (SOZ) [8]. Using the “Epileptogenicity Map” function on Brainstorm software (Version: 3.231027), we quantified the level of increase in fast activity (between 60 to 200 Hz) from the time window immediately before (−20s to 0s) to after seizure onset (0s to 20s) in all SEEG contacts using a statistical parametric mapping procedure implemented in Brainstorm. Accordingly, the level of power in the high-frequency band was quantified over 2s-long sliding time windows (no overlap). This gave us topographic maps on the brain surface indicating how much increase in the fast activity each area experienced upon seizure onset as quantified by t value.

Second, to track the potential flow of epileptogenic activities across the brain we used a simplified version of Granger-based connectivity analysis which has successfully tracked signals across the brain [27]. Here, we included the whole signal without filtering or feature extraction (i.e., broadband) which included slow shifting signals as well as fast activity (Figure 3B). Specifically, we used a partial correlation metric to evaluate the contribution of activity from one area to another:

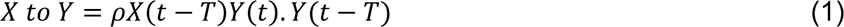

where *ρX*(*t* − *T*)*Y*(*t*). *Y*(*t* − *T*) indicates the correlation between past activity on contact *X* to present activity on contact *Y*, while partialling out the influence of past activity on contact *Y*. *X*(*t*) and *Y*(*t*) are activities at time *t* in areas *X* and *Y*, respectively, and *T* is the time/sample delay considered between two areas (here *T* = 100*ms*). In our implementation of the method the signals from two areas along a 1s window (*t* = *t*0, …, *t*0 − 1000*ms*) were used for calculating the correlation (i.e., 200 signal samples were used across pairs of areas). As the partial correlation equation in (1) was asymmetric, we also calculated the opposite flow to quantify the flow from *Y* to *X* contacts and then reported their difference as the dominant flow of information across pairs of contacts. We calculated the signal flow on every 5^th^ sample (5ms sampling period) along the pre to post seizure onset window (−10 to 10s). This gave us a time-resolved connectivity measure quantifying the flow of signal across the sampled areas upon seizure onset. For this analysis, we used the same SEEG contacts that were found to produce the maximum number of spikes interictally, which also overlapped with the contacts that showed the largest increase in fast activity upon seizure onset. We set the inter-contact delay (*T*) to 100ms consistently across all analysed contacts as this gave us the highest partial correlations on average across all pairs of analysed contacts. It is of note that, as opposed to conventional connectivity analyses, where the connectivity between all contacts are evaluated, we only analysed connectivity between pairs of four contacts targeting the right anterior (N) and posterior (X) cingulate, right amygdala (A) and right mesiotemporal pole (TP), which produced the maximal spiking in the interictal period.

#### Statistical analyses

The significance of epileptogenicity maps were evaluated in Brainstorm software using statistical parameter mapping at a threshold of *α* = 0.01 and corrected for false discovery ratio for multiple comparisons across time, electrode contacts and frequencies. This resulted in t-values. t-values above 1.96 are generally considered to indicate a significant effect (meaning increase in fast activity power from baseline in this work).

As in our previous work [29], we used a Bayes Factor analysis for statistical inference of signal flow upon seizure onset [45]. We used standard rules of thumb for interpreting levels of evidence [12,37]: Bayes factors above 6 and below 1/6 were interpreted as evidence for the alternative and null hypotheses, respectively. We considered the Bayes factors which fell between 1/6 and 6 as insufficient evidence either way.

To evaluate the evidence for the null and alternative hypotheses of at-chance and above-chance signal flow, respectively, we compared the signal flow on every time point and the signal flow obtained from the null distribution. To obtain the null distribution of signal flows, we shuffled the samples of signal *X* a thousand times and calculated the signal flow from *X* to *Y* and reverse, calculated the null signal flow and compared it against the true signal flow. For that, we performed an unpaired Bayes factor t-test for alternative (i.e., difference from chance; H1) and the null (i.e., no difference from chance; H0) hypotheses.

To evaluate the evidence for the null and alternative hypotheses of non-different and different from zero inter-area delay, respectively, we compared the true inter-area delay vector with a vector of the same size but only including zero values.

The priors for all Bayes factor analyses were determined based on Jeffrey-Zellner-Siow priors [25,54] which are from the Cauchy distribution based on the effect size that is initially calculated in the algorithm using t-test [45].

#### Data availability

Images showing seizure semiology are available upon request to the authors.

## Results

### HDEEG, MEG, fMRI

No ictal activity was recorded during the non-invasive recording session (HDEEG alone, HDEEG-MEG, HDEEG-fMRI). Interictal ESI results were, however, consistent across the three sequential acquisition sessions with the early phase source activity identifying a candidate generator at the right anterior frontal lobe, adjacent to the supero-medial resection margin (Figure 2A). MSI localisation was stable at all discharge phases at the right anterior cingulate cortex but was infero-medial to the early-phase ESI localisation (Figure 2B).

**Figure 2.**
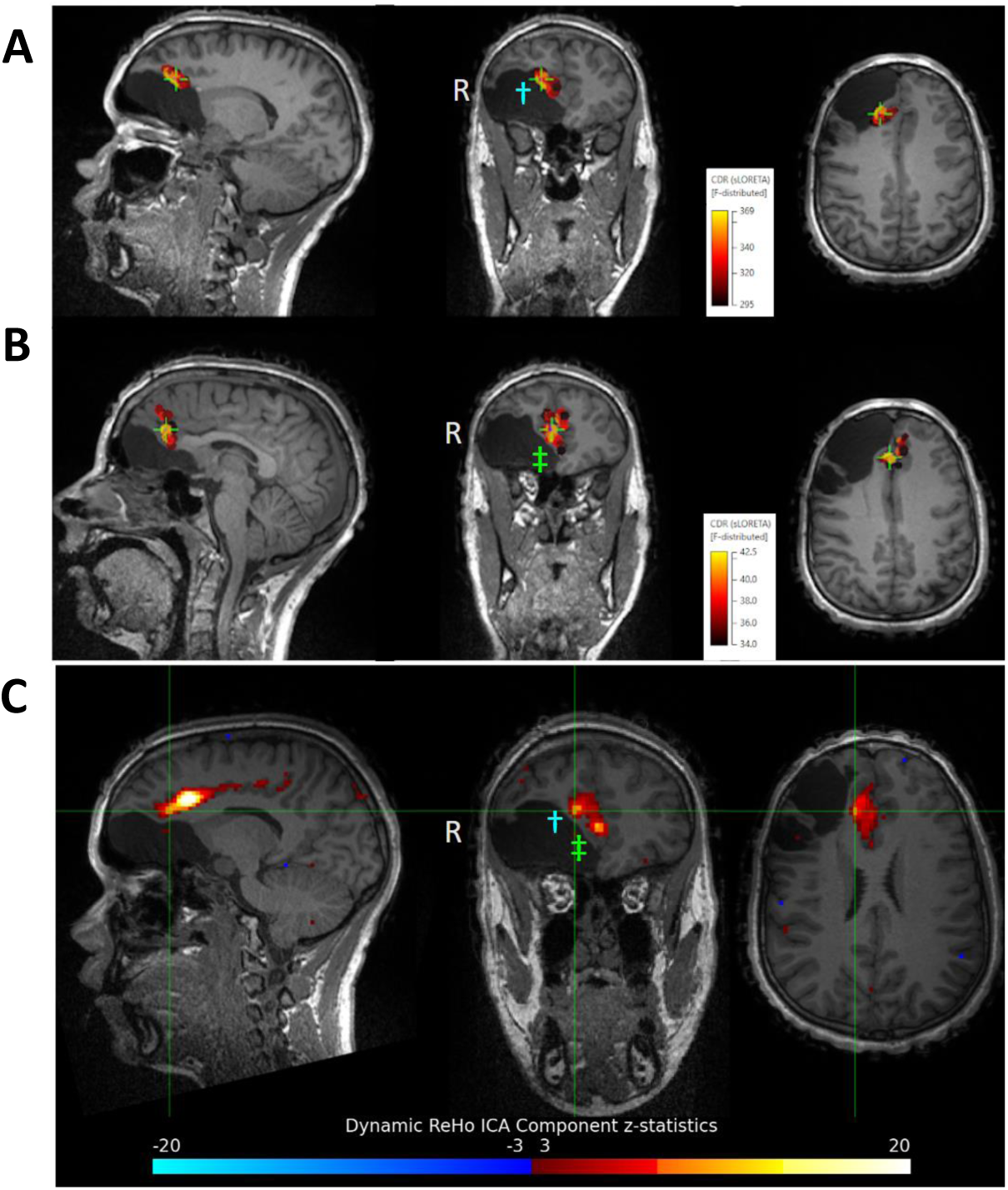
Key non-invasive multimodal imaging results for early phase ESI (**A**), early phase MSI (**B**) and ReHo fMRI (**C**). ESI and MSI solutions were based on averaged interictal discharges carrying the highest SNR values (n=12) during the HDEEG-MEG co-acquisition. ESI and MSI solutions sit at the posterior margin of the prior resection at the right frontal lobe, with MSI inferomedial to the ESI current density result. Note that the broader ReHo fMRI solution co-localises to both ESI and MSI solutions at its anterior extent (ESI co-localisation single dagger blue, MSI co-localisation double dagger green), while its posterior extent involves the anterior cingulate and mid-cingulate junction. Based on the superior temporal resolution of ESI and MSI over fMRI, we hypothesised that the primary EZ generator involved the anterior-most region of the right anterior cingulate while the posterior-most anterior cingulate and middle cingulate regions, which were also incriminated by ReHo fMRI, were more likely expressions of recruited or propagated activity. ESI results were internally consistent across recording sessions (HDEEG alone, HDEEG-MEG, and HDEEG-fMRI).

Routine ICA modelling of denoised fMRI data highlighted BOLD results that were consistent with both the ESI and MSI early onsets, as well as delineating a broader region of the right cingulate cortex that extended posteriorly to the mid-cingulate cortical boundary. With two subregions showing stronger activation hotspots from ICA. Further exploration from the ReHo analysis more clearly identified local connectivity strength of activation, consistent with the ESI and MSI results but uncovering a prominent activation posteriorly along the line of the anterior cingulate towards the right mid-cingulate junction (Figure 2C). Details of additional fMRI analyses including EEG-informed GLM analysis and EEG-naïve Independent Component Analysis are presented in Supplementary Figure 1.

### SEEG

Sample recordings from the interictal and ictal time windows are shown in Figure 3. The patient had consistent and stereotyped patterns of interictal activities over time with increases in the prevalence of spikes in right anterior cingulate area (initial contacts of N) from day 1 to day 5 with decreasing ASMs. We also observed very consistent patterns of ictal activity as reflected in slow-wave shifts and low-voltage fast activity starting in right anterior cingulate area followed by other areas.

**Figure 3.**
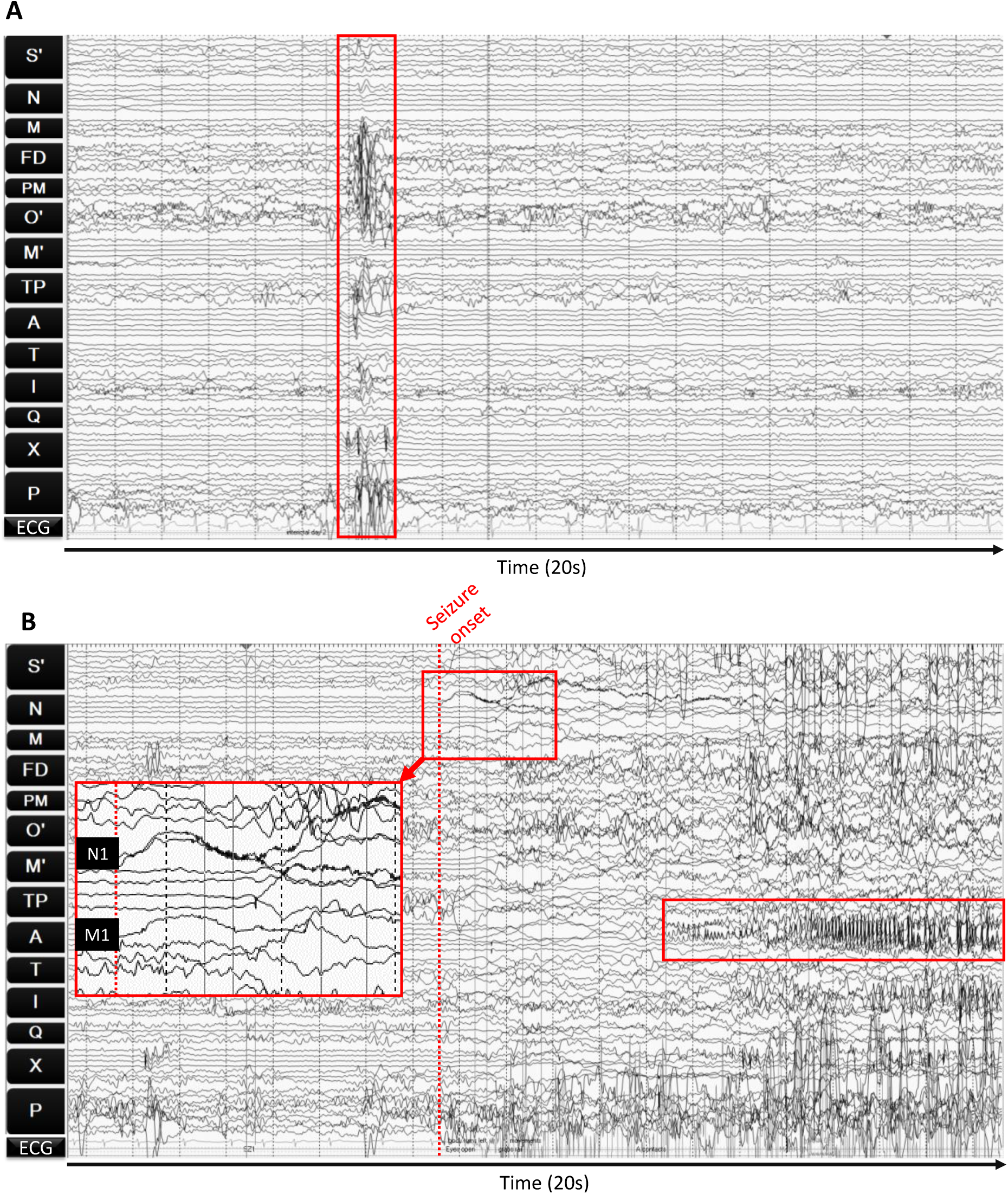
Sample interictal (**A**) and ictal (**B**) data with epileptiform activities indicated by red boxes. **A**, Interictal data shows a combination of spiking and HFOs in the indicated times which are more pronounced in amplitude in the first contacts of electrodes N (R anterior cingulate), throughout FD (R mid frontal gyrus), PM (R mid frontal gyrus), TP (mesiotemporal pole), I (mid frontal gyrus), X (R posterior cingulate) and P (R superior parietal lobe) electrodes. **B**, Ictal activities start by slow wave low-voltage fast activity in initial contacts of electrodes N and M (anterior to mid-cingulate cortices, inset), and TP (mesiotemporal pole) as well as X (posterior cingulate cortex), which then spread to other areas clearly reflected in high-voltage discharges in electrode A (amygdala). This electrographic pattern was highly stereotyped across all seizures.

#### Interictal analysis

Interictal spike detection showed that spikes were most prevalent in right anterior (N) and posterior (X) cingulate, right amygdala (A) and right mesiotemporal pole (TP) (Figure 4A). All the mentioned areas produced the maximum number of spikes on day 1 and 5 except that right anterior cingulate ranked a top spiking site on the last day of recording when the number of seizures were also maximal (Supplementary Figure 2). This introduced a brain-wide structure as the irritative network involving cingulate and mesiotemporal structures.

**Figure 4.**
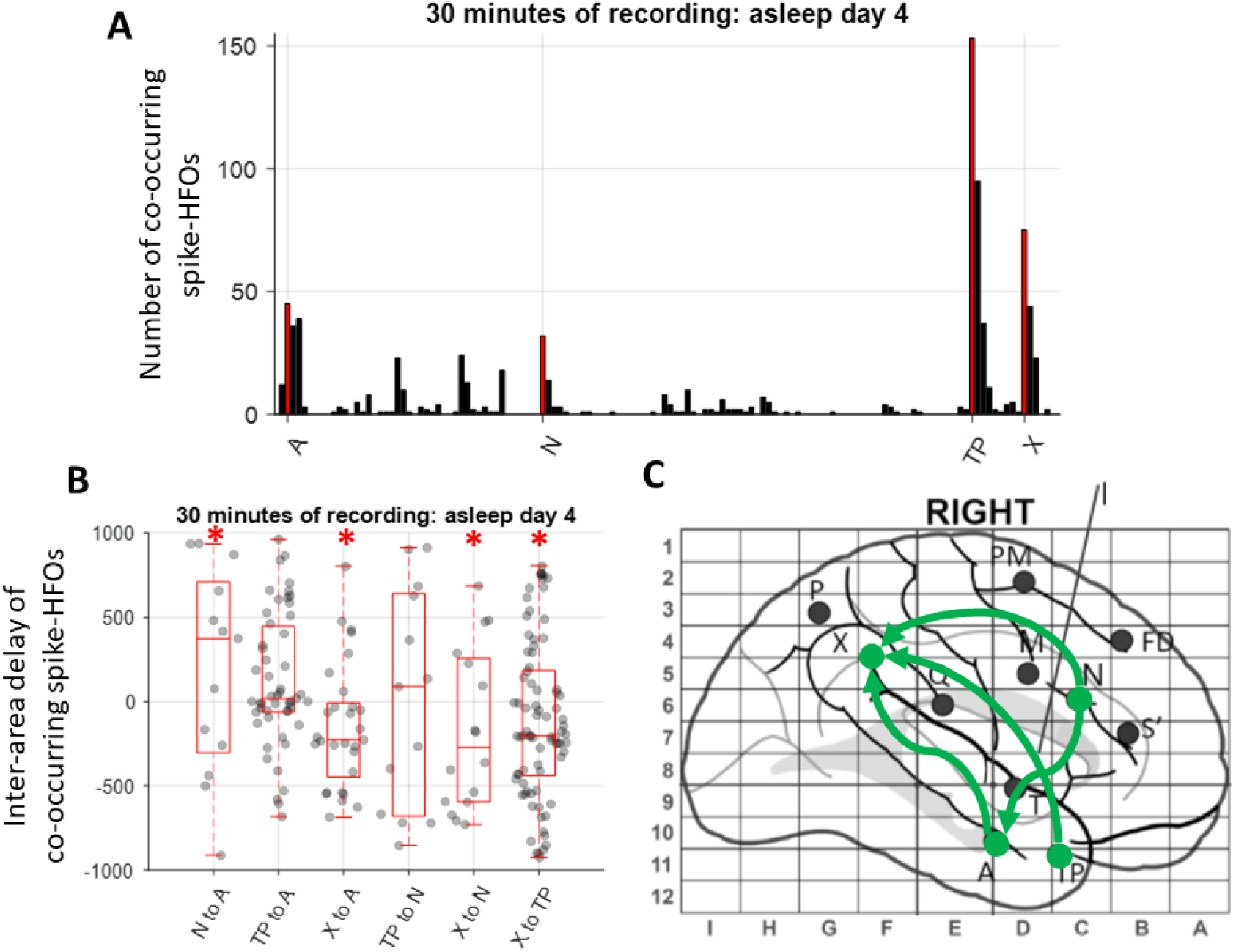
Interictal signal count (**A**), inter-area spike delay (**B**) and the propagation map (**C**). **A**, the number of spikes which were detected in each area is shown with the target contacts indicated in red. **B**, the time delay between the spikes which co-occurred within 1s of each other across all possible pairs of target electrode contacts indicated in **A.** Box plots show the distribution of data, its quartiles and median and whiskers indicate the maximum and minimum of the data. Asterisks indicate the contact pairs across which the delays were different from zero (BF > 6). **C**, the electrode implantation map with potential spike propagations indicated in green. Electrodes targeted left and right anterior cingulate (S’), right anterior cingulate (N), right mid cingulate (M), right frontal superior gyrus (FD), right pre-secondary motor area (PM), left rectus gyrus (O’), left mid cingulate (M’), right mesial temporal pole (TP), right amygdala (A), right inferior insula (T), right dorsal anterior insula (I), right posterior insula (Q), right posterior cingulate (X) and right precuneus (P).

In order to check the potential propagation of spikes between those areas, we extracted the time of spike across the four contacts. The time delays between all co-occurring spikes across all possible pairs of these brain areas are plotted in Figure 4B. Results showed that spikes predominantly appeared earlier in right anterior cingulate than the right amygdala and right posterior cingulate and they appeared earlier in right temporal pole and amygdala than the posterior cingulate (asterisks indicate the contacts across which the delay was different from zero; BF > 6). This suggests that spikes seem to predominantly initiate in the right anterior cingulate and propagate to right mesial temporal pole, amygdala and the posterior cingulate (Figure 4C).

#### Ictal analysis

We obtained epileptogenicity maps over time which quantified the relative increase in the power of fast activity (between 60 to 200 Hz) upon seizure onset across the brain areas sampled by implanted electrodes (Figure 5). These results represent the average of the 9 seizures. We observed that, upon seizure onset (1-3s relative to onset), right anterior and mid cingulate areas (including N and M electrode contacts), showed the highest increase in the power of fast activity. This was followed (3-5s) by many other areas including right and left dorsolateral, temporal, and parietal cortices.

**Figure 5.**
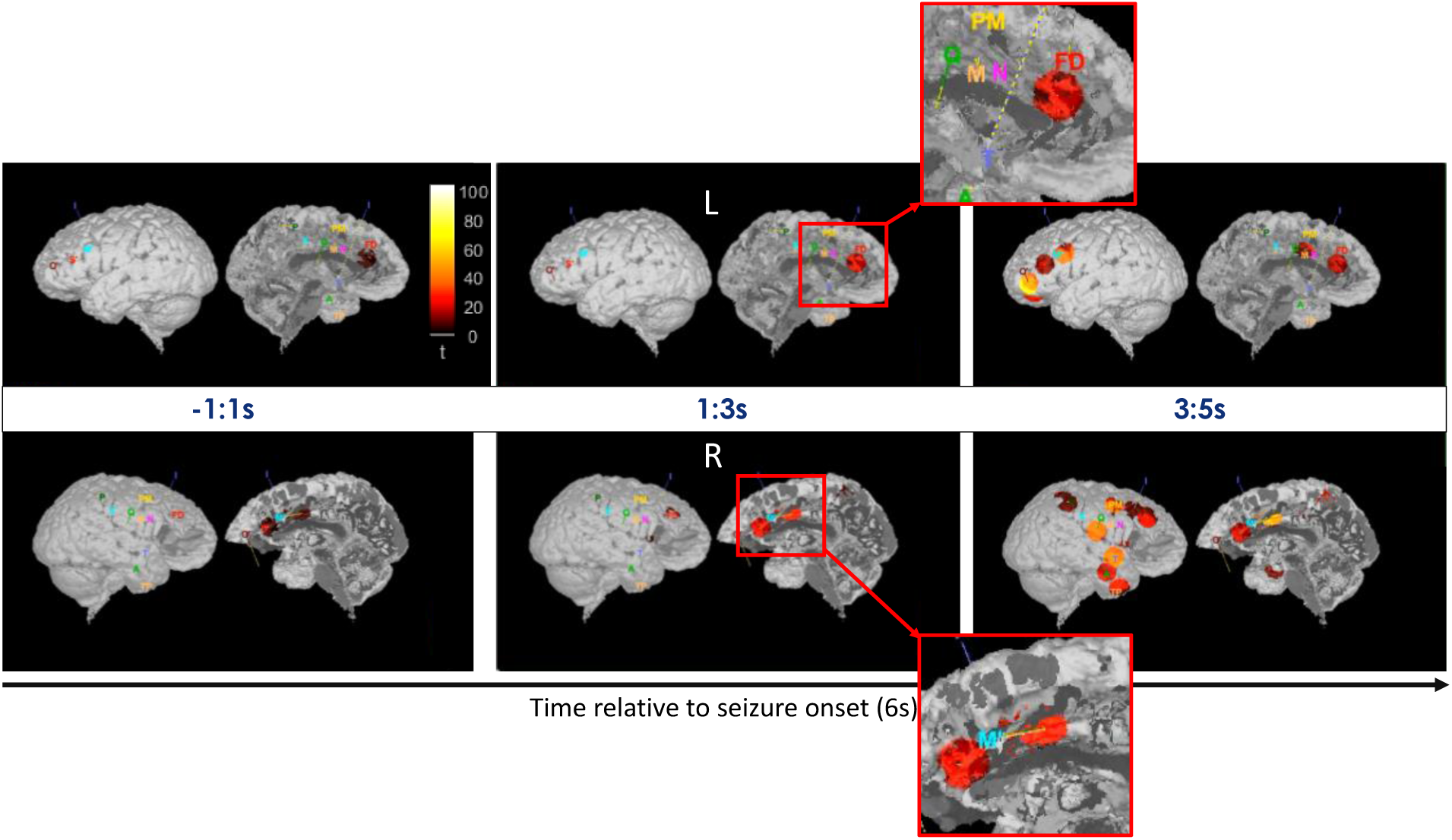
Epileptogenicity maps indicating the level of increase in fast activity (60−200 Hz) across the brain in 2s sliding time windows relative to the baseline before seizure onset (−20 to 0s). Brighter colours indicate higher increase in the power of fast activity suggesting that right anterior to mid cingulate cortices were the first areas to show the increase in high-frequency power (middle column). The area of the activity around each electrode contact is fixed, and the colour changes with the level of increase in fast activity. L: left hemisphere; R: right hemisphere.

The epileptogenicity maps suggest that the fast activity started from the anterior mid-cingulate area and propagated to involve other areas, notably mesial temporal, insular, lateral frontal and posterior cortex. To quantify how the seizure activity flowed across the brain we applied a Granger-based connectivity analysis and tracked the signals between right anterior and posterior cingulate, right amygdala and right temporal pole. These areas were indicated by the interictal spiking activity and also showed the highest increase in fast activity upon seizure onset. It is of note that, as opposed to the epileptogenicity map analysis, here we included the broad-band signals rather than the fast activity only. Around the seizure onset time (−1 to +2s) there was strong evidence (Bayes Factor > 6) for flow of signals from right anterior cingulate cortex to the right mesial temporal pole (Figure 6A).

**Figure 6.**
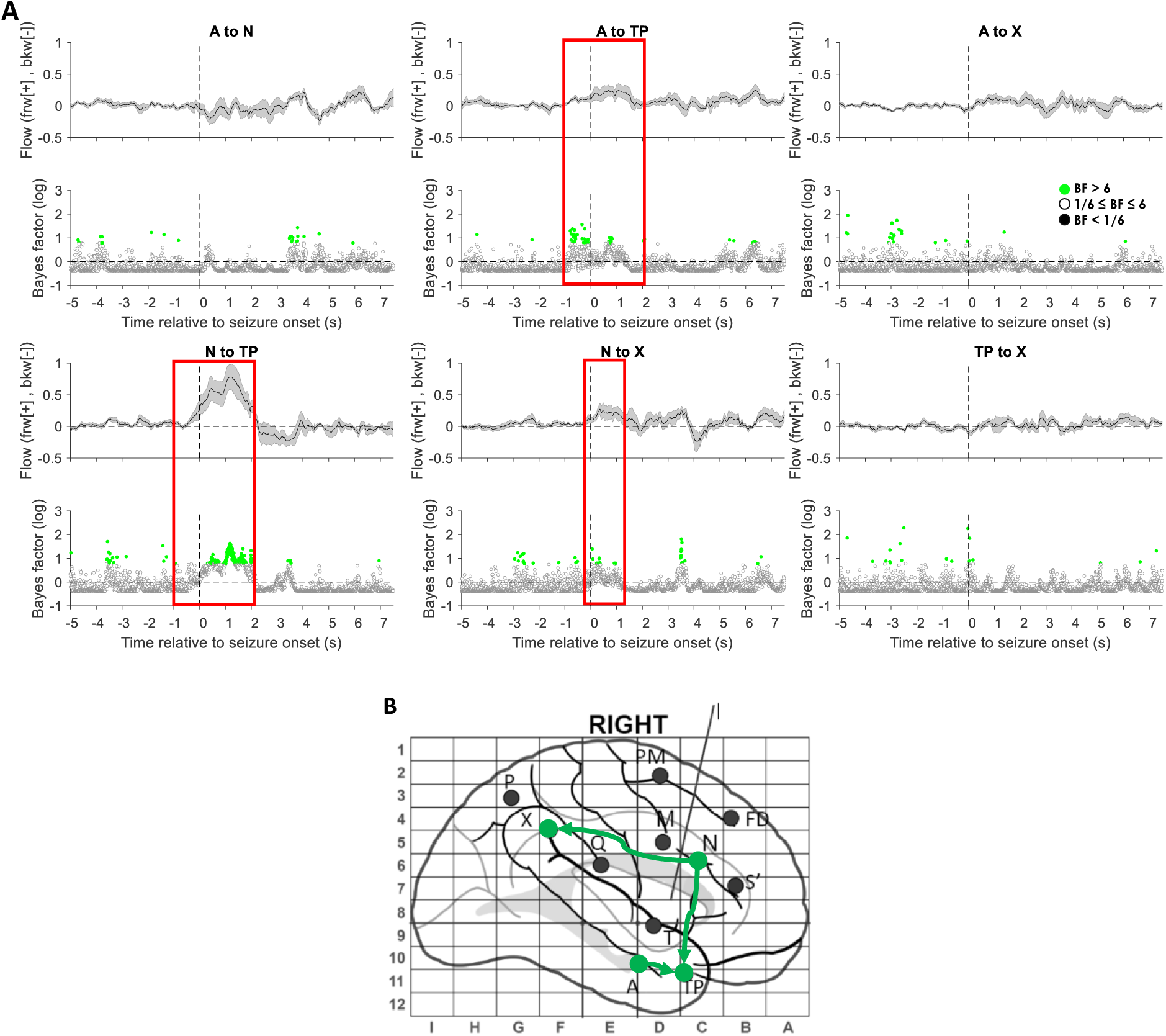
Granger-based connectivity analysis of signal flow during seizures. The flow of signals across pairs of brain areas are evaluated on every 5ms. **A**, each panel indicates the direction and value of the flow averaged across all nine seizures (shading: standard error over seizures). Positive flow reflects feed-forward flow (in the direction noted in the title of the panel) and negative flow reflects feedback flow (opposite to the direction indicated in the title). Bottom panels show the result of Bayesian evidence for and against above-chance flow. Bayes Factors (BF) above 6 are considered strong evidence for the flow of information across pairs of areas. Red rectangles indicate the time points when there was consistent evidence for above-chance flow of information. **B**, the electrode implantation map with potential signal propagations indicated in green based on **A**. Electrodes targeted left and right anterior cingulate (S’), right anterior cingulate (N), right mid cingulate (M), right frontal superior gyrus (FD), right pre-secondary motor area (PM), left rectus gyrus (O’), left mid cingulate (M’), right mesial temporal pole (TP), right amygdala (A), right inferior insula (T), right dorsal anterior insula (I), right posterior insula (Q), right posterior cingulate (X) and right precuneus (P).

There were also time points with strong evidence (BF > 6) for signals flowing from the right amygdala to right mesial temporal pole. There were also sporadic time points with strong evidence (BF > 6) for signals flowing from right anterior to posterior cingulate cortices. These results are summarised in the graphical panel in Figure 6B. Together, the connectivity results of ictal data suggested that the seizure signals initiated in right anterior mid-cingulate areas and then propagated to the right posterior cingulate and mesial temporal areas.

## Discussion

We performed a multi-modal analysis to localise the EZ in a patient with hyperkinetic seizures due to MRI-negative, focal, drug-resistant epilepsy. This case was challenging clinically because of complex semiology, no evident MRI epileptogenic lesion and persistent frequent disabling hyperkinetic seizures despite two previous right frontal resections, one of which had shown evidence of FCD type 2A. As such, the decision to pursue further presurgical investigation required careful analyses of non-invasive data in order to refine hypotheses to be tested using SEEG, thus shaping implantation strategy, in a setting where standard clinical data alone did not allow for sufficiently detailed hypotheses to be formulated, notably because of complex semiology and absence of MRI lesion (apart from prior surgical cavity). The ensemble of non-invasive results helped to reinforce that the right frontal lobe, and in particular right mesial frontal regions, showed most abnormalities. Our novel signal analyses applied to a combination of HDEEG, MEG, fMRI and SEEG datasets provided concordant results indicating that the primary EZ was most likely in the right prefrontal region, maximally centred upon the right anterior and mid-cingulate area.

The multi-modally concordant results of this study not only provided evidence for the value of multimodal analysis in localisation of EZ, but we also introduced several novel quantitative approaches for EZ localisation.

### HDEEG, MEG, fMRI

Our novel single session, sequential HDEEG-MEG and HDEEG-fMRI recordings and results demonstrate consistent inter-modality localisation of the putative EZ to the region of the right anterior cingulate gyrus. This information was critical in guiding the placement of the SEEG electrodes, which subsequently validated our non-invasively acquired findings. These results are also in line with our previous work that demonstrates the importance of looking at earlier phase components closer to the region of take-off for the source analysis of interictal (and ictal) discharges (as opposed to limiting ESI and MSI to the later mid-upswing and peak components) [4,42]. Contemporary source imaging approaches still tend to focus on these later time-points for EZ mapping despite the risk that the solutions, while carrying higher SNR values, are more likely to be contaminated by the effects of extensive cortical recruitment and propagation [51]. The rapidity with which cortical propagation can occur for interictal events, which is less well appreciated than for ictal events, is clearly evidenced by the subsequent SEEG findings in this patient. Although the HDEEG-MEG and HDEEG-fMRI recordings did not capture ictal activity in our patient, our previous work has highlighted the commonality of ictal and interictal patterns of signal origin and propagation when the whole epoch from discharge onset to peak is submitted to source imaging [42]. This observation finds further support in the follow-on SEEG interictal and ictal analyses, which both strongly implicated a common generator in the region of the right anterior cingulate cortex.

The central finding of the EEG-informed GLM models highlighted a stronger frontal correlation with BOLD activations, however the EEG-naïve ICA approaches uncovered consistent broader cingulate involvement extending posteriorly to the mid-cingulate region. The transformation of the fMRI data into dynamic measures of regional homogeneity (local connectivity) clarified the spatial relationships of activated cortex that included those identified by ESI and MSI. The power of the multimodal acquisitions enabled us to cross validate features identified independently using different parameters to confirm spatial concordance of solutions from both EEG-naïve and EEG-informed models.

Taken together, the ESI, MSI, and fMRI results suggested that either a complex source or complex multi-nodal network was responsible for interictal source generation. The benefits of the spatial resolution offered by fMRI remained hampered by the slower haemodynamic response relative to the higher EEG and MEG temporal resolutions. While this does limit our ESI-MSI to fMRI cross modality interpretation in discriminating whether a generator is uniformly distributed or focal, the extent of the activated cortical regions provided a clearer hypothesis for SEEG exploration. Our complementary non-invasive modalities suggested that the primary EZ generator localised to the right anterior frontal regions at the posterior resection margin and anterior cingulate (as indicated by the early phase ESI and MSI) before recruitment of the posterior-aspect of the right anterior cingulate and mid-cingulate areas (as indicated by the fMRI features).

### SEEG

Our spike delay analysis in SEEG is novel and builds on the recent studies which looked at the timing of spikes across recording contacts to localise the EZ [11,47]. Specifically, one study investigated the fine-grained timing and direction of interictal and ictal discharges using microelectrode grids and showed that interictal discharges are travelling waves that traverse the same path as seizure discharges [47]. After confirming a consistent temporal ordering of discharges in interictal and ictal windows, another study developed a novel source localisation method based on wave propagation, which successfully localised the EZ [11]. Our spike delay analysis builds on these previous studies and shows that the direction of spike propagation in the interictal period is consistent with the flow of broad-band signals in the ictal period (i.e., both appearing first in the anterior cingulate cortices). The generalisation of interictal spikes to broad-band ictal power is important because the seizure activity generally consists of spatiotemporally complex broad-band activities rather than only ictal spiking, albeit with evidence that the key fast activity at seizure onset is typically narrow gamma band, as evidenced in Chauvel and colleagues’ “fingerprint” analyses [21] and also highlighted in recent work [10]. The separation of ictal spikes from non-spike activity imposes difficulties for analysis and may always involve some subjective decisions on labelling of spikes.

Importantly, both interictal spiking and ictal seizure patterns in SEEG showed an outward propagation from the anterior cingulate area, which we considered to be the EZ core, and propagated to the secondary involved areas (referred to as propagation zone previously [35]. While these results are intuitive, the full characterisation of the relationships between the epileptogenic patterns in the two time windows need further research as the literature is not fully consistent. On one hand, we have recently shown that some global features of neural activities, such as beta-band power and gamma-band inter-channel coherence (a measure of connectivity), can have localising power in both interictal and ictal periods [32]. To test the generalisability of patterns over time we trained the classifiers using the data from one time window (e.g., interictal) and tested them to localise the EZ in the other time window (e.g., ictal) and found the same features to be informative and provide above-chance localisation performance. On the other hand, there are studies which have shown that while the signals flow towards the seizure onset zone in the interictal period, consistent with the source-sink theory of EZ [22], the direction is dominantly opposite in the ictal period, suggesting distinct patterns of signal flow in the two windows [26]. This seems at odds with our results and those reviewed above showing the consistent direction of spikes and broad-band signals in the interictal and ictal periods, respectively.

This discrepancy can be explained by the fact that these studies have used distinct features of neural activity (e.g., spikes vs. broad-band signal) and distinct measures of directed connectivity (e.g., spike timing vs. coherence) each of which may affect the results. Future studies that evaluate multiple features of signals in isolation [28] or combination [31] along with more rigorous connectivity methods [30] on the same dataset may provide a more comprehensive insight into the dominant direction and consistency of signal flow in interictal and ictal periods and whether that can localise the EZ.

These analyses complement and go beyond the clinician’s standard visual analysis, not only by quantifying signals related to interictal and ictal pathophysiologic activity, but also indicating their directionality, which helps to reinforce the separation of EZ from propagation zones. This is a particularly challenging aspect in frontal lobe seizures explored on SEEG due to their known propensity for rapid and widespread propagation related to intrinsic properties of frontal lobe connectivity [3]. In the present case these analyses helped to confirm for example that, while the extra-frontal inter-ictal spikes were overall more abundant than the mesial frontal spikes, the temporal relation of their occurrence was in favour of a primary role of the anterior mid-cingulate region (i.e., same region as maximal seizure onset fast activity). Likewise, the seizure onset analyses of directionality of gamma band activity helped to show an antero-posterior spread in the cingulate that was maximal at a distance from the previous resection margin. This helped to define surgical strategy in a way that would not have been possible using for example intraoperative electrocorticography at the previous resection margin.

#### Limitation

We recognise that the main limitation of this work is that it has been presented here as a single case report. While the application of the proposed methods to additional patients is needed to establish their generalisability, the main contributions of this work are twofold. Our approach helps qualify preoperative targets non-invasively and provides more confidence in the use of SEEG in characterising and validating markers of complex extended epileptogenic networks leading to more successful surgical outcomes. This method presents a natural augmentation and paradigm-shift from the original solid SEEG investigation to a multi-modal approach by leveraging more recent technological advancements to broaden both temporal and spatial resolutions (and biomarker dimensions).

## Conclusion

Multi-modal HDEEG-MEG-fMRI non-invasive data analyses can optimise patient selection for SEEG and can help to guide SEEG implantation strategy and surgical decision-making in complex cases. The recording of brain activity across these three modalities within a single session (when the patient is more likely to be in the same “brain-state”) can also be achieved, allowing ESI, MSI, and fMRI results to more fully characterise the extent of the EZ. Invasive quantitative analyses demonstrated overlap with our combined non-invasive findings, lending confidence to our hypothesis for the likely EZ and opening the way for a more definitive surgical solution for our patient. As non-invasive multi-modal investigation paradigms are validated by SEEG localisation of EZ and satisfactory surgical outcomes, it may become possible to rely more on non-invasive investigations for cerebral localisation in selected cases. Larger case series with robust comparison of multi-modal data in different clinical sub-groups will be necessary to better understand the predictive value of these investigations.

## Data Availability

All data produced in the present study are available upon reasonable request to the authors. Images showing seizure semiology are available upon request to the authors.

## Acknowledgements

We would like to thank all members of the Mater Epilepsy team, Brisbane including Dr Jason Papacostas, Neurosurgeon. We thank Prof Simon Harvey’s team (Royal Children’s Hospital, Melbourne) and Prof Patrick Chauvel (Cleveland Clinic Foundation) for helpful clinical case discussions. We thank Mater Foundation and Mater Research Institute for supporting this study. We would also like to acknowledge the facilities and scientific and technical assistance of the National Imaging Facility, a National Collaborative Research Infrastructure Strategy (NCRIS) capability, at the Swinburne Neuroimaging Facility, Swinburne University of Technology.

## Supplementary Materials

**Supplementary Table 1.**
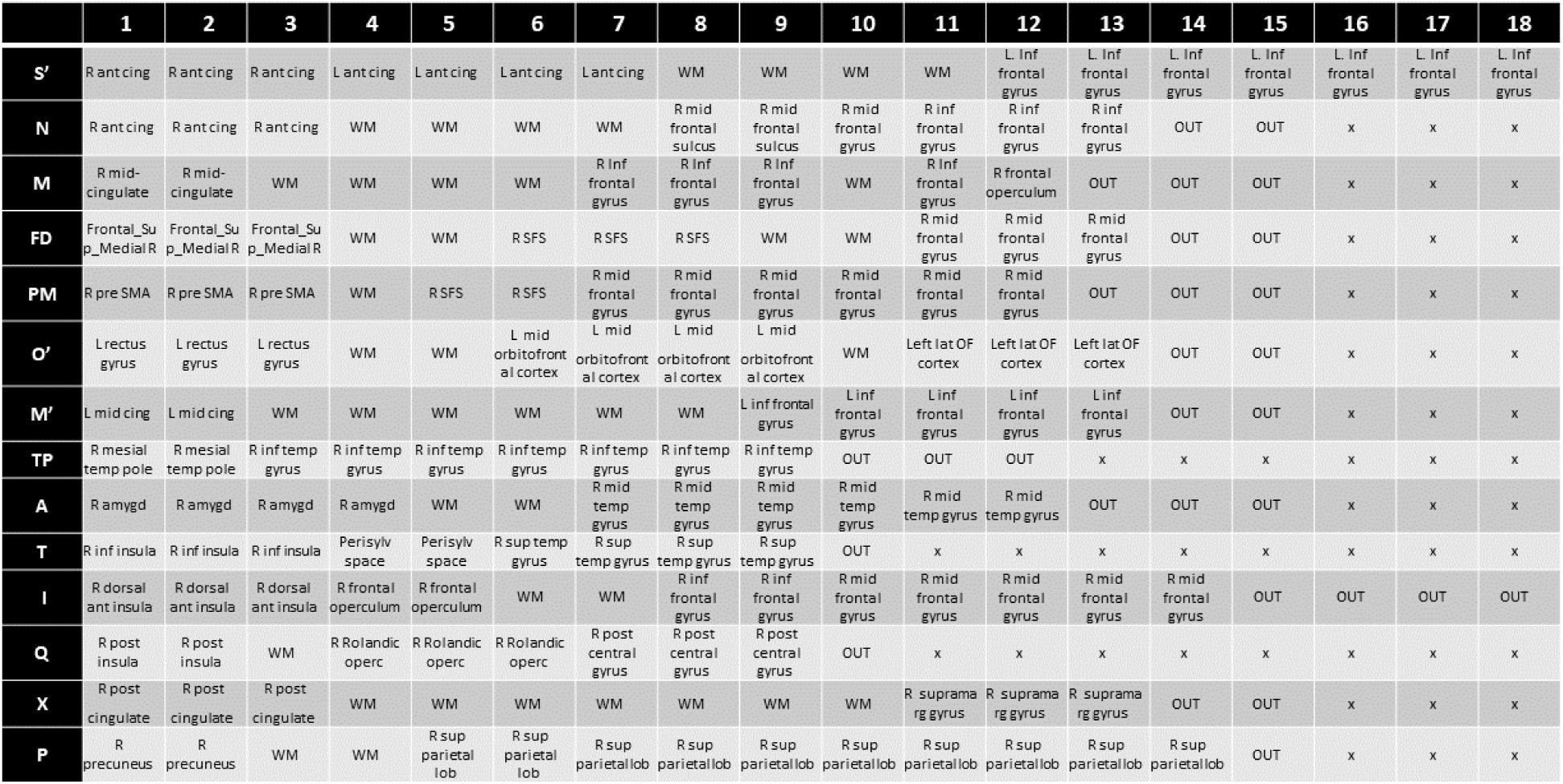
Location of each SEEG electrode contact in the brain according to Hammers 95 atlas. Rows indicate electrodes and columns indicate contact number with lower numbers being the most internal in the brain. x: no contact, WM: white matter, OUT: contact outside of the brain.

**Supplementary figure 1.**
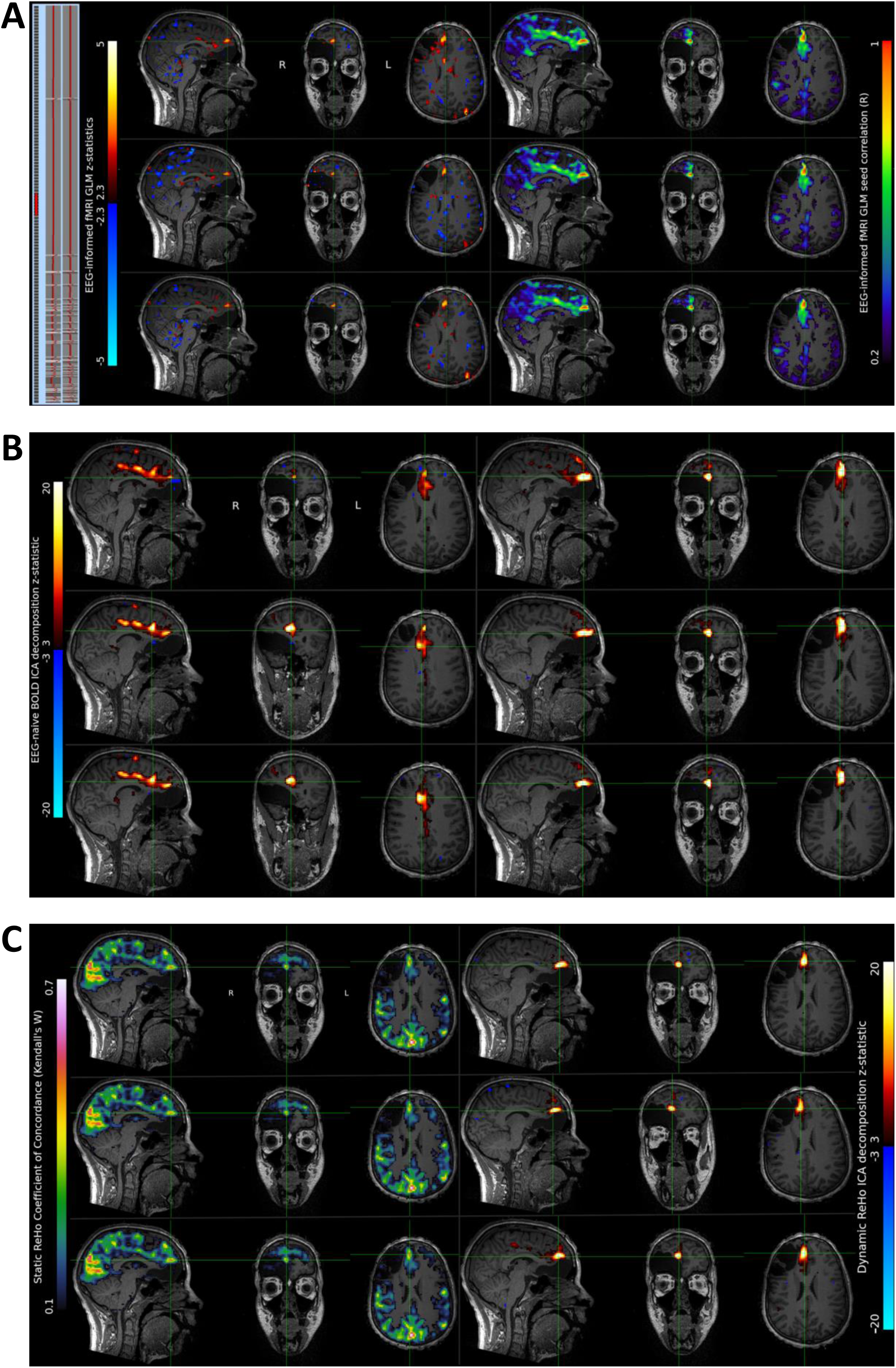
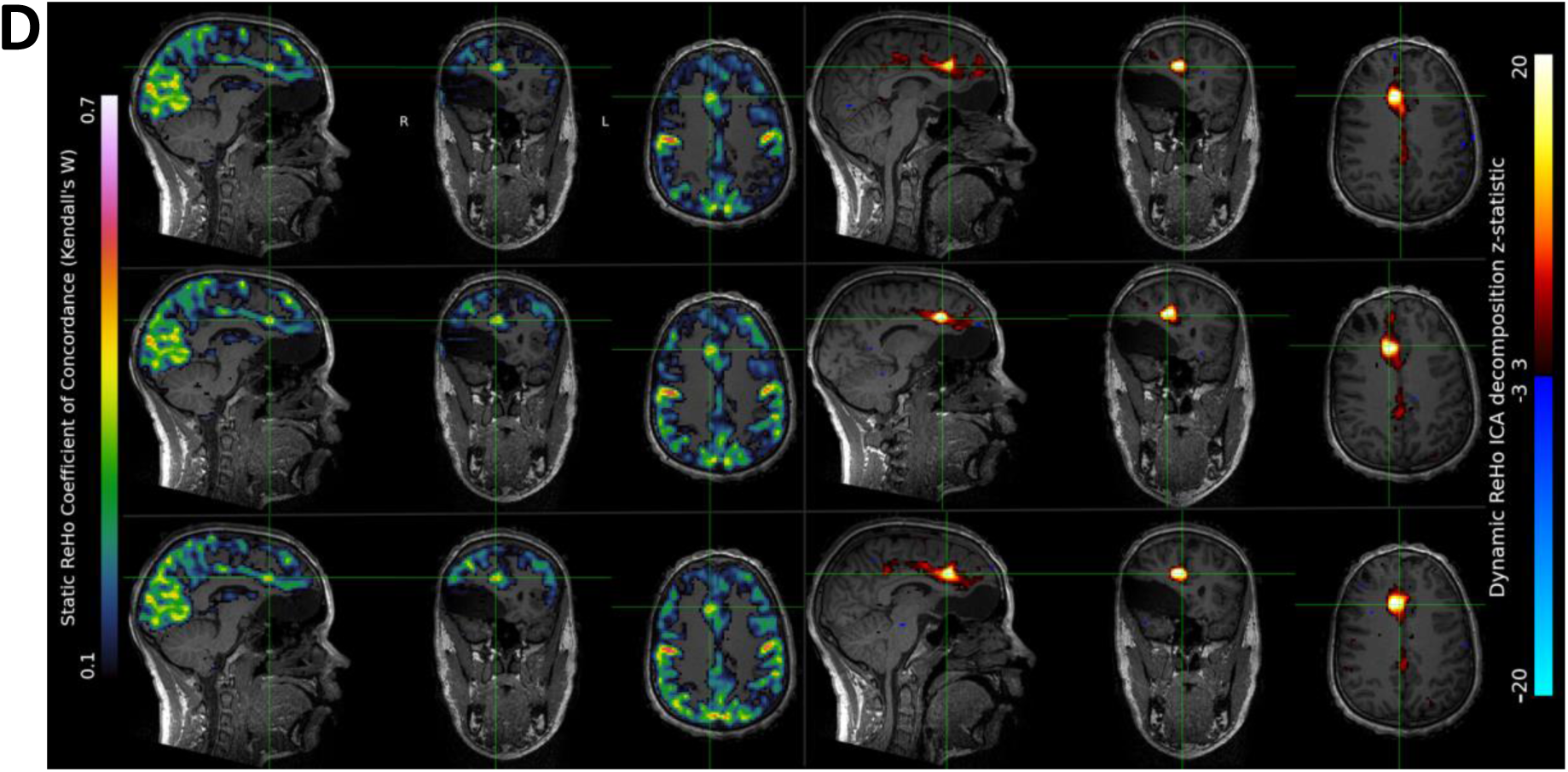
Additional fMRI analyses. **A**, an EEG-informed GLM analysis from manually identified interictal epileptiform discharges using denoised fMRI data (regressor of epileptiform event timings and their temporal derivative shown at far left) highlights maximal response at the right mesial frontal region. Three-row panels (Upper, 24.37ms; Middle, 38.14ms; Lower, 51.91ms) represent sagittal, coronal and axial views of results from independent preprocessing and analysis of simultaneous acquired Multi-echo multi-band fMRI from these increasing Echo time (TE), respectively. The maximal right mesial frontal voxel from each analysis was used to generate a Pearson’s R seed correlation map, highlighting consistent connectivity patterns involving the cingulate cortex in these same three planes. **B**, two EZ candidates for the epileptogenic zone identified from EEG-naïve Independent Component Analysis of the denoised multi-echo multi-band fMRI data acquired during simultaneous high-density EEG-fMRI. The three echo time fMRI datasets (Panels: Upper 24.37, Middle 38.14 and Lower 51.91ms) captured consistent spatial ICA maps (shown in sagittal, coronal and axial) involving the right cingulate cortex and right mesial frontal cortex, respectively, across each panel, highlighting minor variation likely from effects of echo dependence on BOLD signal. **C**, right mesial frontal EZ candidate identified from static Regional Homogeneity (ReHo) connectivity map (using Kendall’s W coefficient of concordance; left sagittal, coronal and axial MRI overlays) and identification of spatial ICA sub-component from dynamic ReHo z-statistical map (right set of orthogonal MRI overlays), across the three independent TE time series panels representing (Upper 24.37, Middle 38.14ms and Lower 51.91ms denoised datasets respectively). The static ReHo map clearly identifies areas of higher local connections commonly associated with visual functional connectivity posteriorly in the occipital cortex, but whole brain thresholding of such measures cannot automatically identify culprit generators for the EZ. The ICA subcomponents from dynamic ReHo do allow for interrogating independent variations in localised functional connectivity. **D**, right anterior cingulate EZ candidate suggested from static Regional Homogeneity (ReHo) connectivity map (using Kendall’s W coefficient of concordance; left sagittal, coronal and axial MRI overlays) and identification of spatial ICA sub-component from dynamic ReHo z-statistical map (right set of orthogonal MRI overlays), across the three independent TE time series panels representing (Upper 24.37, Middle 38.14ms and Lower 51.91ms datasets respectively). The static ReHo map identifies a discrete area of higher local connections at the right anterior cingulate cortex, but the dynamic ReHo ICA analysis offers evidence for the extent of the nodal region involved in generating coherent activity, including reference to slightly weaker statistical involvement with the mesial frontal and mid-cingulate cortex. Such patterns may help discern complex nodal interactions from non-invasive modalities for subsequent verification by invasive modalities such as SEEG.

**Supplementary figure 2.**
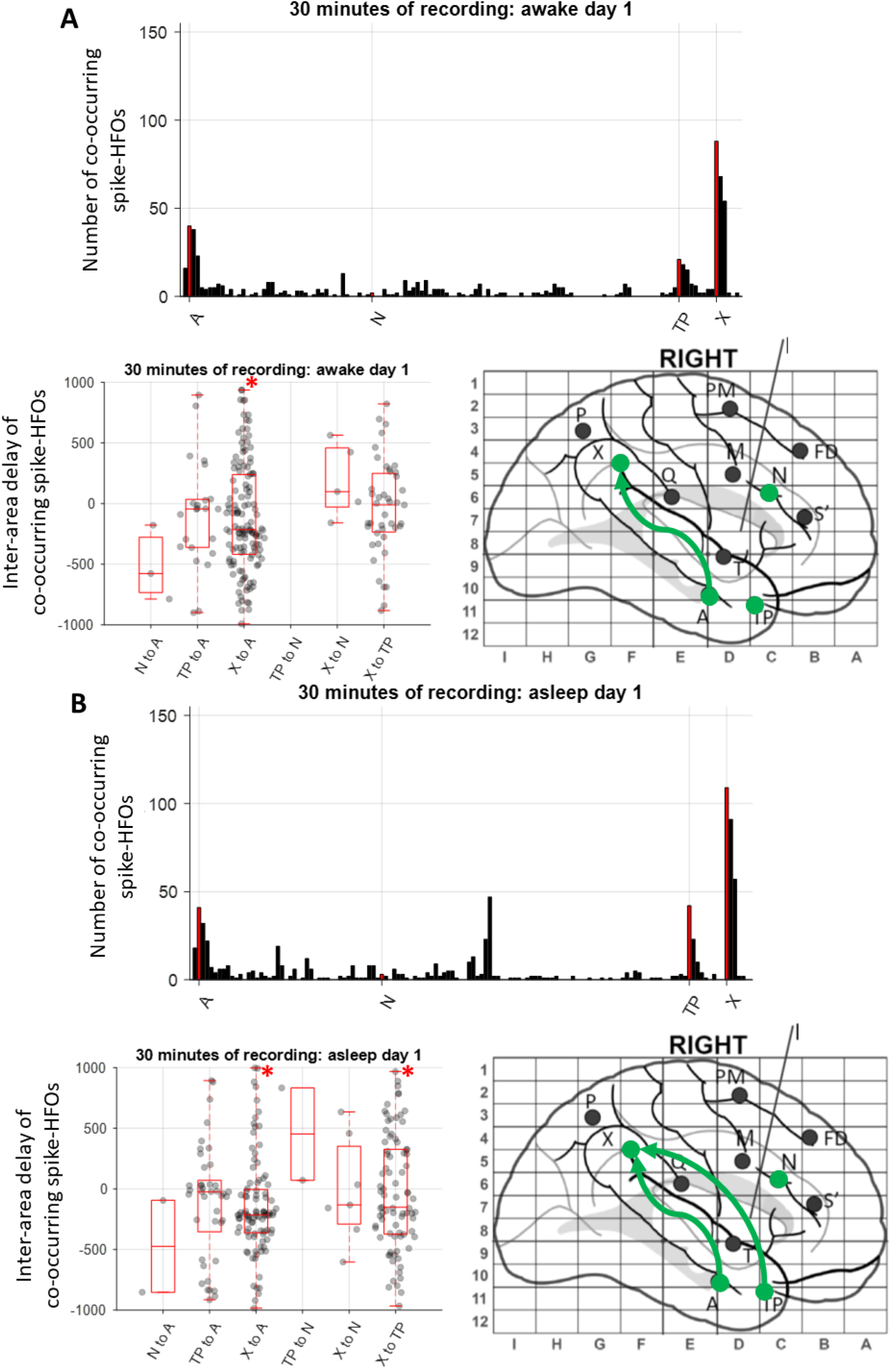
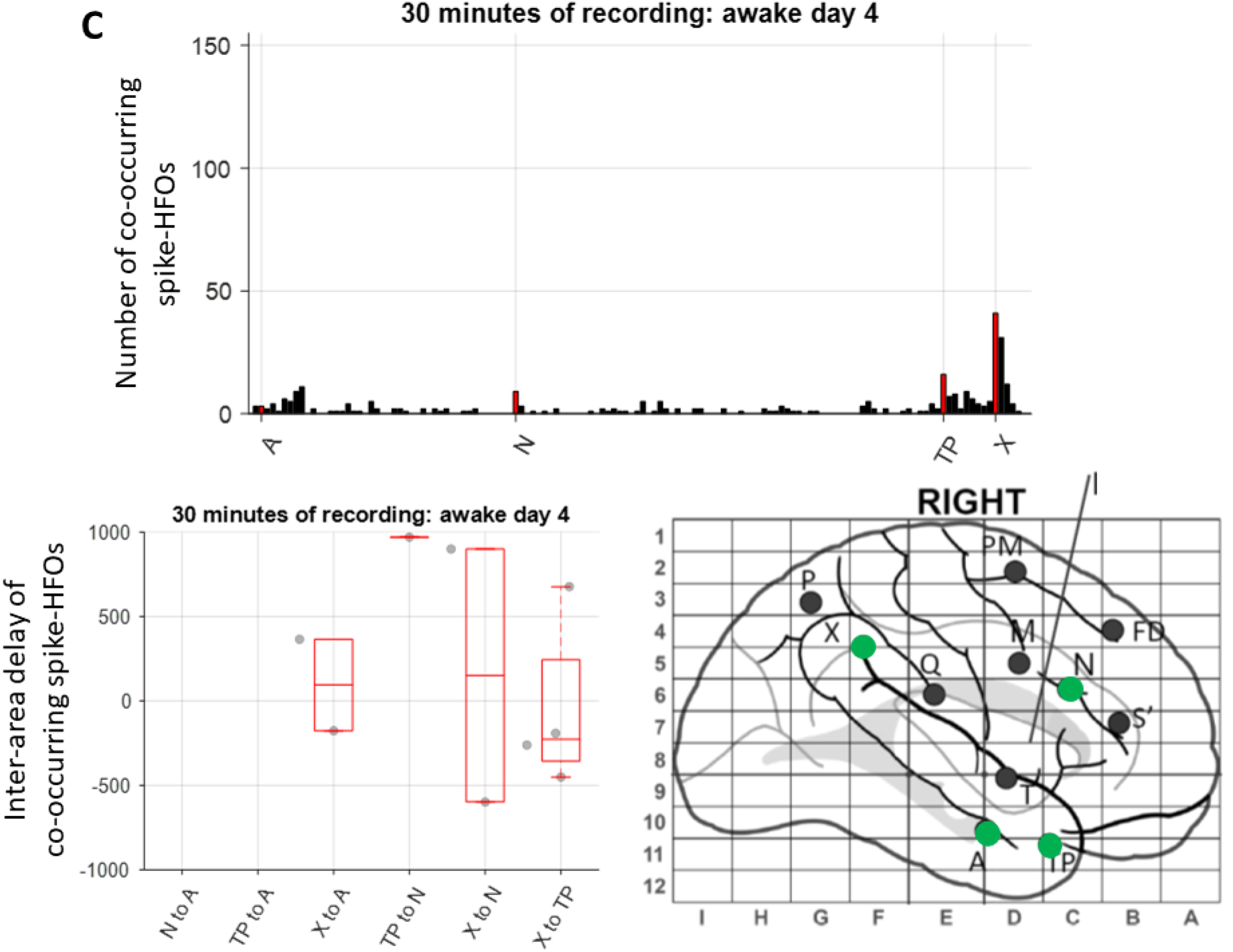
Interictal signal count, inter-area spike delay and the propagation map. **A** shows the results from day 1 in awake state, **B** shows the results from day 1 in asleep state and **C** shows the results from day 4 in awake state. Top panels, the number of spikes which were detected in each area is shown with the target contacts indicated in red. Bottom left panels, the time delay between the spikes which co-occurred within 1s of each other across all possible pairs of target electrode contacts. Box plots show the distribution of data, its quartiles and median and whiskers indicate the maximum and minimum of the data. Asterisks indicate the pair-wise delays which were different from zero (BF>6). Bottom right panels, summary maps indicating potential direction of co-occurring spikes across contacts for which the delay was different from zero. Electrodes targeted left and right anterior cingulate (S’), right anterior cingulate (N), right mid cingulate (M), right frontal superior gyrus (FD), right pre-secondary motor area (PM), left rectus gyrus (O’), left mid cingulate (M’), right mesial temporal pole (TP), right amygdala (A), right inferior insula (T), right dorsal anterior insula (I), right posterior insula (Q), right posterior cingulate (X) and right precuneus (P).

